# Association among income loss, financial strain and depressive symptoms during COVID-19: evidence from two longitudinal studies

**DOI:** 10.1101/2020.09.15.20195339

**Authors:** N Hertz-Palmor, TM Moore, D Gothelf, GE DiDomenico, I Dekel, DM Greenberg, LA Brown, N Matalon, E Visoki, LK White, MH Himes, M Schwartz-Lifshitz, R Gross, RC Gur, RE Gur, IM Pessach, R Barzilay

## Abstract

**Background:** The COVID-19 pandemic has major ramifications for global health and the economy, with growing concerns about economic recession and implications for mental health. Here we investigated the associations between COVID-19 pandemic-related income loss with financial strain and mental health trajectories over a 1-month course.

**Methods:** Two independent studies were conducted in the U.S and in Israel at the beginning of the outbreak (March-April 2020, T1; *N* = 4 171) and at a 1-month follow-up (T2; *N* = 1 559). Mixed-effects models were applied to assess associations among COVID-19-related income loss, financial strain, and pandemic-related worries about health, with anxiety and depression, controlling for multiple covariates including pre-COVID-19 income.

**Findings:** In both studies, income loss and financial strain were associated with greater depressive symptoms at T1, above and beyond T1 anxiety, worries about health, and pre-COVID-19 income. Worsening of income loss was associated with exacerbation of depression at T2 in both studies. Worsening of subjective financial strain was associated with exacerbation of depression at T2 in one study (US).

**Interpretation:** Income loss and financial strain were uniquely associated with depressive symptoms and the exacerbation of symptoms over time, above and beyond pandemic-related anxiety. Considering the painful dilemma of lockdown versus reopening, with the tradeoff between public health and economic wellbeing, our findings provide evidence that the economic impact of COVID-19 has negative implications for mental health.

**Funding:** This study was supported by grants from the National Institute of Mental Health, the US-Israel Binational Science Foundation, Foundation Dora and Kirsh Foundation.

## Introduction

The COVID-19 pandemic has major ramifications for global health^1^. To reduce virus spread, many countries have ordered strict measures to impose social distancing^2^. While such preventive measures are effective in reducing transmission, their economic costs are overwhelming^3,–5^. In April 2020, unemployment in the US reached 14·7%, a 10·3 percent increase compared to the previous month, with 15·9 million newly unemployed workers. This is the steepest one-month increase since 1948^3^. Similar trends have been observed in other areas of the worldsup^4^. The World Bank has estimated that by the end of 2020, international global gross domestic product (GDP) will decline by up to 5·2%, the worst global recession in 80 years^5^. Behind the numbers, reduced GDP means greater unemployment, which is a major risk factor for worsened mental health^6^..Unemployment increases psychological strain as, besides the loss of income, it is accompanied with loss of social contact, status and sense of competence^7^. Thus, the economic climate in mid-2020 poses an imminent threat to mental health deterioration^8^, adding to the health-related stress invoked by the pandemic. Some evidence from the 2007 economic crisis suggests that economic stress is specifically associated with seeking mental health help due to depression^9^.

Recent findings indicate that the current COVID-19 crisis is no exception, with its economic impact hitting hard on global mental health. In China, high levels of anxiety and depression were associated with worries about income, job, study or inability to pay loans^10^. In a large nationally representative study conducted in the UK, people who were unemployed or had no income (e.g. were full-time students) during the pandemic were more distressed than those employed. Furthermore, people who lost their job during the pandemic expressed greater increase in distress than individuals who were already unemployed before COVID-19^11^. This finding might reflect the link between the sharp decrease in financial security and deterioration in mental health experienced by many workers; however, this study did not address the specificity of financial concerns to mental health beyond other concerns such as COVID-19 health-related worries^11^. A study from the U.S reported higher rate of depressive symptoms during the pandemic compared with pre-pandemic, highlighting low household income and savings as risk factors for depression; however this study did not compare financial stressors with health-related stressors^12^. In a study from Switzerland, the impact of financial concerns on internalizing symptoms (depression, anxiety, self-injury and suicidal ideation) among young adults (*N* = 768, mean age 22) exceeded that of health-related concerns^13^. This finding is in line with a report that young people are less physically threatened by the virus, but are more vulnerable to losing their job^14^. In times of an ongoing debate about whether economic considerations should be prioritized over health^15^, there is need for data to clarify the link between finances and mental health and their trajectories during COVID-19^1,16^.

We recently described a crowdsourcing platform (covid19resilience.org) that collected data on COVID-19 related stress (worries) and mental health in population primarily composed of US and Israel participants sampled during the acute pandemic outbreak^17^. In the current research, we investigated the specific association between self-reported financial consequences of COVID-19 and mental health (anxiety and depression) during the pandemic, over and above the impact of health-related concerns. We further analyzed data from an independent study conducted in Israel. In each study we investigated (1) cross sectional associations between financial hardship and concurrent mental health and (2) mental health trajectories over a month during the pandemic. We hypothesized that loss of income (objective financial hardship) and perceptions of financial strain (the subjective feeling of economic well being^18^) would be associated with increased anxiety and depression symptoms and would predict the exacerbation of symptoms over time above and beyond health related COVID-19 concerns. We further tested whether financial stressors would be uniquely associated with depression, above and beyond their effect on anxiety symptoms.

## Methods

### Study 1

#### Participants and procedure

Participants were ascertained through a crowdsourcing website (https://covid19resilience.org/) that collected data on COVID-19 related stress (worries), resilience and mental health^17^. At the end of the survey, participants received personalized feedback on their responses. The feedback was meant to incentivize participants to complete the survey carefully. Participants were offered the opportunity to leave their email address and be contacted for follow up surveying. The study was advertised through, 1) the researchers’ social networks, including emails to colleagues around the world; 2) social media; 3) the University of Pennsylvania and Children’s Hospital of Philadelphia internal notifications and websites; and 4) organizational mailing lists. The survey was available in English and Hebrew. The results presented are based on data collected from April 6^th^ to May 5^th^, 2020 for the baseline cross sectional analysis (T1); Participants who left their email address in T1 and consented to be re-contacted received an email inviting them to participate in a longitudinal survey between May 12^th^ and June 21^st^ (T2). Participation required responders to provide online consent. The study was approved by the Institutional Review Board of the University of Pennsylvania.

### Measures

Anxiety and depression were measured using the Generalized Anxiety Disorder-7 (GAD-7) questionnaire^19^ and Patient Health Questionnaire-2 (PHQ-2)^20^, respectively. COVID-19-related worries (self-contracting COVID-19, family contracting COVID-19, financial burden due to COVID-19) were measured on a 5-point Likert-type scale (from not at all to a great deal). Participants were also asked whether they had lost their job or whether their pay/hours were reduced since the beginning of the outbreak (collapsed into a binary income loss measure [yes/no]).

### Statistical analysis

#### Cross sectional models

Linear mixed-effects models were applied to investigate associations of financial hardship with mental health (anxiety and depression symptoms) as the dependent variable. In the first model, we considered income loss (yes/no) as the independent variable. In the second model, we considered the COVID-19 related financial strain (stress/worry) as the independent variable, contrasted against the stress of self-contracting COVID-19, which served as the reference variable for pandemic-related worries. This model also included the stress of a family member contracting COVID-19. Symptom type (anxiety or depression) was addressed as within-person repeated measure. Models included multiple potential confounders: age, gender, pre-COVID-19 annual income (in USD (ordinal measure)), relationship status, living alone, country of origin and date of survey completion.

#### Longitudinal models

For the longitudinal analysis we used linear mixed-effects models with the symptoms as the key dependent variable, where change over time could be observed due to the longitudinal nature of the data. Model 1 included dynamics (change) in income loss from T1 to T2. Change in income loss consisted of four possible values (1-0, 0-0, 1-1, 0-1; 0=no loss; 1=loss), where income loss at T2 was considered as deterioration in financial situation (even if already reported at T1, due to expected regression to the mean). Model 2 considered change in worries between T1 and T2 and was calculated as the difference in responses between time-points. Symptom type (anxiety or depression) was addressed as within-person repeated measure. Covariates were the same as cross-sectional models in addition to duration between T1 and T2.

All analyses were performed using the lmerTest package^21^ in R.

### Study 2

#### Participants and procedure

Participants were adult Israelis recruited through a survey link distributed among the researchers’ social media network (WhatsApp contacts and in public groups on Facebook). Data were collected between March 18^th^ and 26^th^ (T1), and again between April 22^nd^ and May 7^th^ (T2, for participants who agreed to be re-contacted for longitudinal surveying through email). All surveys were conducted in Hebrew. Participation required providing online consent. The study was approved by the Institutional Review Board of Sheba Medical Center.

#### Measures

We screened for anxiety and depression with the Hebrew version of the Patient-Reported Outcomes Measurement Information System (PROMIS) Anxiety and Depression modules ^22,23^. COVID-19-related stressors (worries) were compiled from questions that have been shown to be pertinent to mental health in previous research on the SARS and N1H1 pandemics^24^, and were measured on a 4-point Likert-type scale (From not at all to always). Sociodemographic questions were the same as in study 1 with the exception of income (from considerably below average to considerably above average) and income loss (from no income loss to extreme income loss) that were measured on a 5-point Likert-type scale.

#### Statistical analysis

We employed similar models as described in study 1 with a few alterations: The key dependent variables in the cross-sectional and longitudinal models were PROMIS scores

(instead of GAD7/PHQ2). Income loss was on a 5-point scale (instead of binary in study 1), therefore change in income loss was also calculated as the difference in responses between T1 and T2.

## Results

### Study 1

#### Sample characteristics

Cross sectional data (T1) was collected from 5 717 participants who completed the resilience survey and the COVID-19 related questions. Of them, 2 904 people completed GAD-7 and PHQ-2 and were included in this study. Participants were mostly female (*n* = 2 259, 77·8%) with mean age of 41·97 (*SD* = 13·55; range 18 to 91). 815 participants (28·1%) were in the top income category (annual income>$150,000), 556 (19·0%) reported they experienced income loss, and 223 (7·7%) endorsed the top category of financial worries due to COVID-19. In the longitudinal survey (T2), data was obtained from 1 318 participants (54·8% response rate out of *n* = 2 404 that were re-contacted after providing their email in T1). Prevalence of females (*n* = 1 077, 81·7%), mean age (*M* = 40·79, *SD* = 13·59), income loss (*n*= 246, 18·7%) and high financial strain (n = 99, 7·5%) resembled T1. Cohort characteristics are presented in **Table 1**.

**Table 1.**
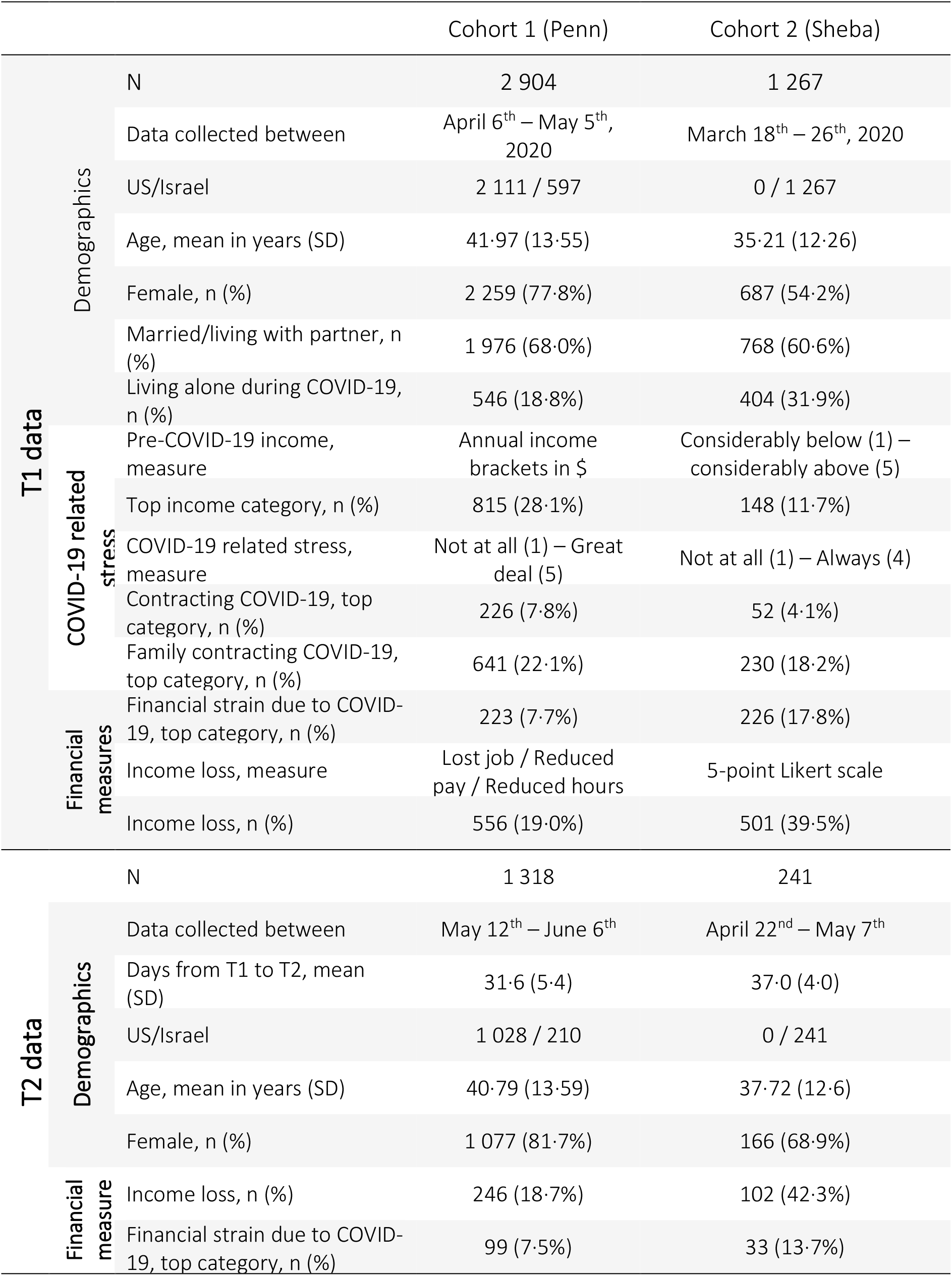
Cohorts’ characteristics

|  |  | Cohort 1 (Penn) | Cohort 2 (Sheba) |
| --- | --- | --- | --- |
| T1 data | N | 2 904 | 1 267 |
|  | Data collected between | April 6<sup>th</sup> – May 5<sup>th</sup>, 2020 | March 18<sup>th</sup> – 26<sup>th</sup>, 2020 |
|  | US/Israel | 2 111 / 597 | 0 / 1 267 |
|  | Age, mean in years (SD) | 41·97 (13·55) | 35·21 (12·26) |
|  | Female, n (%) | 2 259 (77·8%) | 687 (54·2%) |
|  | Married/living with partner, n (%) | 1 976 (68·0%) | 768 (60·6%) |
|  | Living alone during COVID-19, n (%) | 546 (18·8%) | 404 (31·9%) |
| | Pre-COVID-19 income, measure | Annual income brackets in \$ | Considerably below (1) – considerably above (5) |
|  | Top income category, n (%) | 815 (28·1%) | 148 (11·7%) |
|  | COVID-19 related stress, measure | Not at all (1) – Great deal (5) | Not at all (1) – Always (4) |
|  | Contracting COVID-19, top category, n (%) | 226 (7·8%) | 52 (4·1%) |
|  | Family contracting COVID-19, top category, n (%) | 641 (22·1%) | 230 (18·2%) |
|  | Financial strain due to COVID-19, top category, n (%) | 223 (7·7%) | 226 (17·8%) |
|  | Income loss, measure | Lost job / Reduced pay / Reduced hours | 5-point Likert scale |
|  | Income loss, n (%) | 556 (19·0%) | 501 (39·5%) |
| T2 data | N | 1 318 | 241 |
|  | Data collected between | May 12<sup>th</sup> – June 6<sup>th</sup> | April 22<sup>nd</sup> – May 7<sup>th</sup> |
|  | Days from T1 to T2, mean (SD) | 31·6 (5·4) | 37·0 (4·0) |
|  | US/Israel | 1 028 / 210 | 0 / 241 |
|  | Age, mean in years (SD) | 40·79 (13·59) | 37·72 (12·6) |
|  | Female, n (%) | 1 077 (81·7%) | 166 (68·9%) |
|  | Income loss, n (%) | 246 (18·7%) | 102 (42·3%) |
|  | Financial strain due to COVID-19, top category, n (%) | 99 (7·5%) | 33 (13·7%) |

#### Cross sectional association of financial factors with mental health

Higher pre-COVID-19 income was negatively associated with overall anxiety and depression (GAD7 and PHQ2) symptom load (*β* = −0.02, *SE* < 0.01, *t (14 070)* = -3.41, *p* < .001). Income loss during COVID-19 (losing job or reduced pay) was associated with greater depressive symptoms, but not anxiety (**Figure 1A**, income loss by symptom type (anxiety/depression) interaction, *p* < .0001) and had no main effect on general (anxiety and depression) symptom load (*p* = .738).

**Figure 1.**
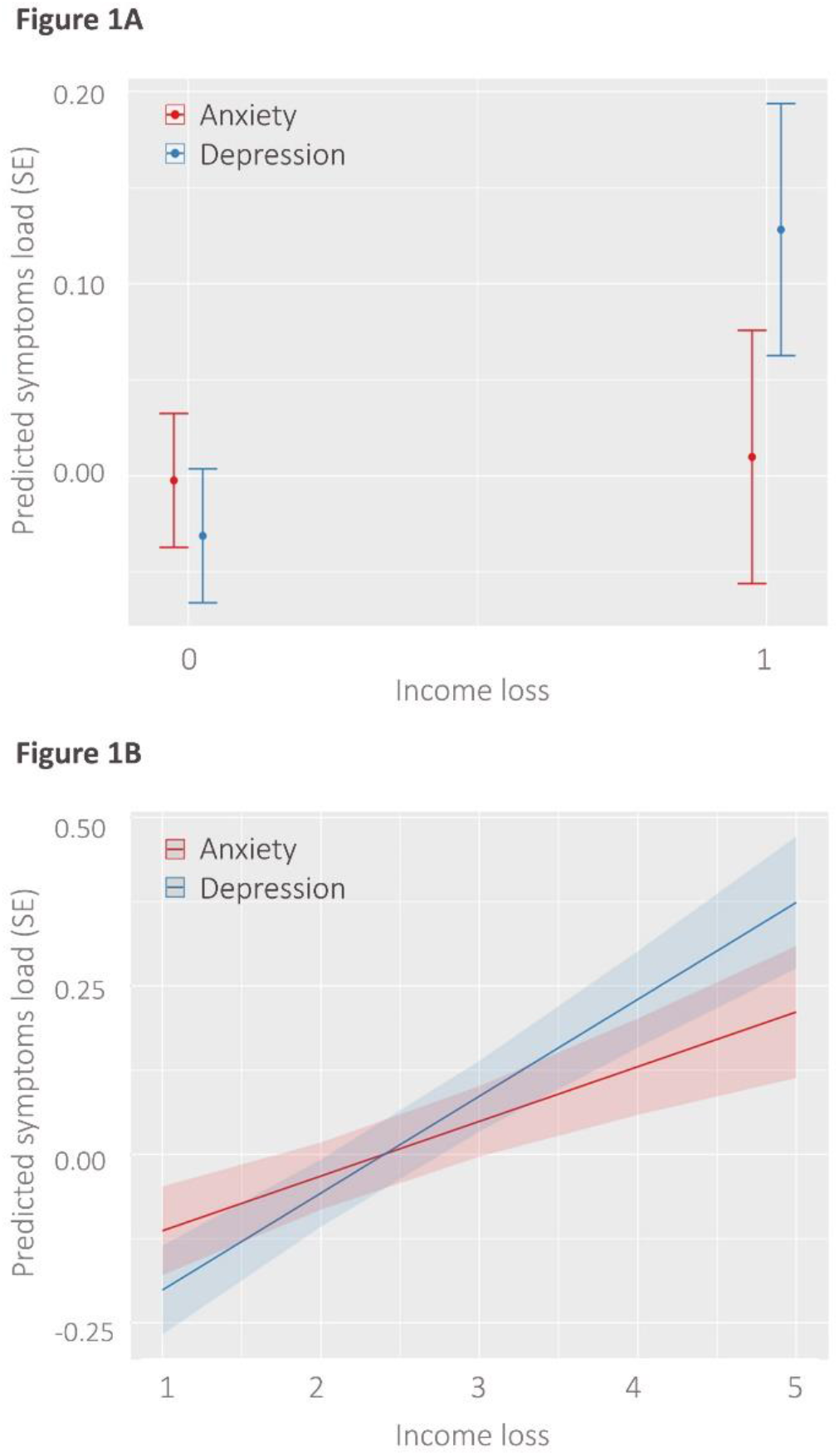
Cross sectional associations between objective hardship (income loss) with anxiety and depression. (A) Associations in Cohort 1; 1 represents losing job\reduced pay\reduced hours. (B) associations in Cohort 2; x-axis represents income loss in a 5-point Likert scale from not at all [1] to extreme income loss [5]. y-axis is standardized (z) symptom score/load.

COVID-19 related worries (self-contracting, family contracting and financial worries) were associated with greater symptom load (*β* = 0.13, *SE* = 0.01, *t (15 280)* = 13.35, *p* < .0001). Financial worries were specifically associated with depressive symptoms (**Table 2**, financial worries by symptom type interaction, *p* < .0001).

**Table 2.** Cross-sectional associations of financial hardship with mental health

|  | <u>Cohort 1</u> |  |  | <u>Cohort 2</u> |  |  |
| --- | --- | --- | --- | --- | --- | --- |
| | Standardized $\beta$<br>(SE) | t | p | Standardized $\beta$<br>(SE) | t | p |
| **<u>Model 1: Objective financial hardship</u>** |  |  |  |  |  |  |
| Income- *main effect on symptoms load (anxiety and depression)* | -0.02 (0.01) | -3.41 | <0.001 | -0.02 (0.02) | -1.37 | 0.168 |
| Income loss- *main effect on symptoms load (anxiety and depression)* | 0.01 (0.04) | 0.33 | 0.738 | 0.08 (0.02) | 4.90 | <0.0001 |
| Specific effect of income loss on depression<sup>#</sup> | 0.14 (0.02) | 8.19 | <0.0001 | 0.06 (0.01) | 8.75 | <0.0001 |
| **<u>Model 2: Subjective financial strain</u>** |  |  |  |  |  |  |
| Overall COVID-19 worries- *main effect on symptoms load (anxiety and depression)* | 0.13 (0.01) | 13.35 | <0.0001 | 0.11 (0.01) | 7.86 | <0.0001 |
| Worries about family contracting COVID-19- *specific effect on Depression<sup>@</sup>* | -0.01 (0.01) | -1.03 | 0.304 | -0.02 (0.02) | -1.19 | 0.232 |
| Worries about finance- *specific effect on Depression<sup>@</sup>* | 0.17 (0.02) | 10.35 | <0.0001 | 0.15 (0.02) | 6.08 | <0.0001 |

Values were derived from linear mixed-effects models with financial hardship (objective in Model 1; subjective in Model 2) as independent variables, and standardized symptoms load (standardized PHQ-2 + GAD-7 scores in Cohort 1, PROMIS Anxiety and Depression scores in Cohort 2) as dependent variable. Models are adjusted for age, sex, relationship status, income and country of origin. Effect sizes are represented with standardized effect coefficients ( $\beta$ ). SE= Standard error.
<sup>#</sup>Modeled as the interaction between income loss and type of symptom domain (anxiety/depression).
<sup>@</sup>Modeled as the interaction between worry type (family contracting COVID-19/financial concern about impact of COVID-19, contrasted against worries to self-contract COVID-19 as reference) and type of symptom domain (anxiety/depression).

#### Longitudinal association of 1-month dynamics in financial factors with mental health trajectories

Income loss at T2 was associated with 1-month increase in general symptoms load (*β* = 0.26). The increase in anxiety symptoms was steeper than that of depression symptoms (**Figure 2A, Table 3**).

**Table 3.** Longitudinal (1-month) association of financial hardship with trajectories of mental health

|  | <u>Cohort 1</u> |  |  | <u>Cohort 2</u> |  |  |
| --- | --- | --- | --- | --- | --- | --- |
|  | Mean (SD) days from T1: 31.6 (5.4) |  |  | Mean (SD) days from T1: 37.0 (4.0) |  |  |
| | Standardized $\beta$<br>(SE) | t | p | Standardized $\beta$<br>(SE) | t | p |
| **<u>Model 1: Trajectory of objective financial hardship</u>** |  |  |  |  |  |  |
| Income loss trajectory- *main effect on increase in symptoms load (anxiety and depression)* | 0.26 (0.02) | 14.66 | <0.0001 | 0.01 (0.05) | 0.23 | 0.817 |
| Income loss trajectory- *specific effect on increase in depression<sup>#</sup>* | -0.07 (0.02) | -4.89 | <0.0001 | 0.07 (0.03) | 2.76 | 0.005 |
| **<u>Model 2: Trajectory of subjective financial strain</u>** |  |  |  |  |  |  |
| Worries trajectory- *main effect on increase in symptoms load (anxiety and depression)* | 0.13 (0.02) | 6.20 | <0.0001 | 0.07 (0.04) | 1.65 | 0.098 |
| Worries about family contracting COVID-19 trajectory- *specific effect on increase in depression<sup>@</sup>* | 0.05 (0.04) | 1.06 | 0.285 | 0.01 (0.09) | -0.03 | 0.975 |
| Worries about finance trajectory- *specific effect on increase in depression<sup>@</sup>* | 0.07 (0.03) | 2.10 | 0.035 | 0.02 (0.08) | 0.32 | 0.741 |

Values were derived from linear mixed-effects models with financial hardship (objective in Model 1; subjective in Model 2) as independent variables, and standardized symptoms load (standardized PHQ-2 + GAD-7 scores in Cohort 1, PROMIS Anxiety and Depression scores in Cohort 2) as dependent variable. Models are adjusted for age, sex, relationship status, income and country of origin. Effect sizes are represented with standardized effect coefficients ( $\beta$ ). SD= Standard deviation. SE= Standard error.
<sup>#</sup>Modeled as the interaction between trajectory of income loss and change in type of symptom domain (anxiety/depression).
<sup>@</sup>Modeled as the interaction between trajectory of change in worry type (family contracting COVID-19/financial concern about impact of COVID-19, contrasted against worries to contract COVID-19) and type of symptom domain (anxiety/depression).

**Figure 2.**
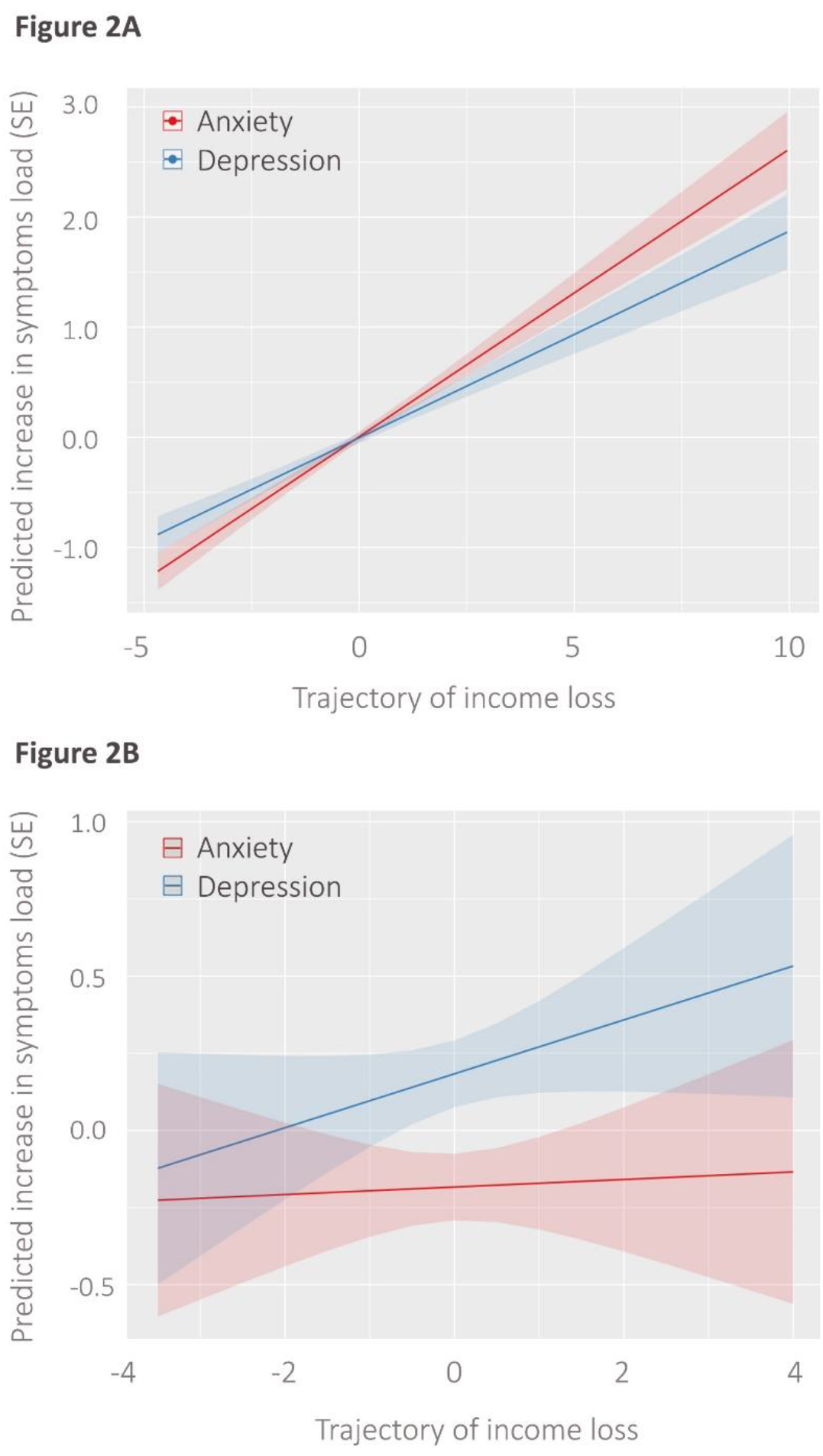
Associations between one-month trajectories of objective financial hardship (income loss) and anxiety and depressive symptoms in longitudinal assessment. (A) Cohort 1 and (B) Cohort 2. X- axis represents change in income loss from T1 to T2, regressed for T1. Negative values on the x-axis indicate that participants reported income loss the first time (T1), and then by the second time (T2), they reported milder income loss. y-axis is standardized (z) symptom load.

Increase in COVID-19 related worries between T1 and T2 was associated with an increase in general symptoms load (*β* = 0.13, *SE* = 0.02, *t (7 176)* = 6.20, *p* < .0001). Increase in financial worries was specifically associated with depressive symptoms (increase in financial worries by symptom type interaction, *p* < .035). Full model statistics are summarized in **Table 3**.

### Study 2

#### Sample characteristics

We conducted similar analysis on a replication cohort from Israel. Of the 1 376 participants that participated in the survey, 1 267 people completed PROMIS and COVID-19 related questions, and were included in this study. Participants were majority female (*n* = 687, 54·2%) with mean age of 35·21 (*SD* = 12·26). 142 participants (11·7%) were in the top income category (reporting their income to be considerably above average), 501 (39·5%) reported that they experienced income loss, and 226 (17·8%) endorsed the top category of financial worries due to COVID-19. In the longitudinal survey, data was obtained from 241 participants (49·7% response rate out of *n* = 485 that were re-contacted). Compared with T1, the follow up sample included higher prevalence of females (*n* = 166, 68·9%) and lower prevalence of people who endorsed the top category for COVID-19 related financial strain (33, 13·7%). Age (*M* = 37·32, *SD* = 12·60) and rates of income loss (*n* = 102, 42·3%) resembled T1. Cohort characteristics are presented in **Table 1**.

#### Cross sectional association of financial factors with mental health

Higher pre-COVID-19 income had no association with general symptoms (PROMIS depression and anxiety) load (*β* = 0.02, *SE* = 0.02, *t (2 192)* = -1.37, *p* = .168). Income loss during COVID-19 was associated with overall higher symptom load (**Table 1**, standardized effect=.08, *p* < .0001). The association of income loss with depression was greater than the association with anxiety (**Figure 1B**, income loss by symptom type interaction, *p* < .0001).

COVID-19 related worries (self-contracting, family contracting and financial worries) were associated with greater symptoms load (*β* = 0.11, *SE* = 0.01, *t (6 535)* = 7.86, *p* < .0001). Financial worries were specifically associated with depressive symptoms (**Table 2**, financial worries by symptom type interaction, *p* < .0001).

#### Longitudinal (1-month) association of dynamics in financial factors with mental health trajectories

A negative trend in financial situation (i.e., worsening of income loss by T2 compared to T1) was associated with 1-month increase in depression (*β* = 0.07, *p* = .005), but had no association with increase in general symptoms load (*p* = .817, **Figure 2B, Table 3**).

COVID-19 related worries had a trend-level effect on increase in general symptoms load (*β* = 0.07, *p* = .098). Increase in financial worries had no specific association with depressive symptoms (increase in financial worries by symptom type interaction, *p* = .741). Full model statistics are summarized in **Table 3**.

## Discussion

Across two independent studies, we found a specific link between financial factors and depression, above and beyond anxiety, which was greater than the association between health-related worries and depression. In cross-sectional analyses of data collected from over 4 000 participants, both loss of income and financial worries had unique effects on depression in the two studies. In longitudinal analyses of ∼1 500 participants, participants from both cohorts who reported a negative trend in income loss reported an increase in depressive symptoms over time. A specific effect of income loss and of financial worries on longitudinal worsening of depressive symptoms, independent of anxiety, was observed in one of two studies. Our findings are in line with data from previous global crises like the 2008 recession that showed a link between financial factors and increase in depression^25^.

The abrupt loss of income (during the pandemic) was associated with overall more anxiety and depressive symptoms, but was associated more strongly with depression than anxiety symptoms, in both cohorts. This dissociation between anxiety and depression may indicate that acute loss of income is a specific risk factor for depression during the pandemic. Our results from Study 2 suggest that recent income loss (measured on a 5-point Likert scale) contributes not only to initial depressive response (data collected at the onset of the pandemic), but may also cause its amplification over time, with a 1-month exacerbation in depressive symptoms associated with worsening in income loss. While we observed an increase in depressive symptoms in Cohort 1 over time in association with income loss, this effect was also observed with worsening of anxiety, even to a larger extent. It is possible that demographic differences between the two cohorts may explain this difference in findings, as Cohort 1 was sampled from a wealthier population that was likely to have more “financial buffering” capacity (i.e., savings, other financial resources beyond salary), and therefore the impact on depression may not have been pronounced. It is also possible that the binary measure that we had used in Cohort 1 to capture income loss (losing job/reduced pay) did not allow sufficient variability in the data to specifically link the worsening of income loss with depression. Our results are in line with recent COVID-19-related data^12^ and with previous findings from economic crises, which associated unemployment and financial difficulties (e.g., mortgage repayment difficulties, evictions, financial shortage ranging from difficulty in paying loans to paying at the supermarket, etc.) with increased depression and help-seeking behaviors^9,25^.

The link we observed between COVID-19 stress (worries) and depression was specific to financial worries. Health-related worries about contracting COVID-19 or family contracting it were associated with general symptoms load, but not depression specifically. This finding controlled for pre-COVID-19 income, suggesting that the objective financial situation only partly explains variability in depressive symptoms, and that worries about the financial situation may be a sensitive marker (red flag) for depressive symptoms during the pandemic. Coupled with the finding that objective income loss is tied to depressive symptoms and that the results replicated across two independent cohorts, our data converge to support the specific link between financial stressors and depressive symptoms. Previous works have shown that financial strain - the subjective stress about financial concerns - is associated with deterioration in mental health, even more than objective inability to meet financial requirements^18,26^.

The urgent need for solid scientific data on the pandemic’s effect on health has spurred concerns regarding validity and generalizability of COVID-19 related publications^27^. That our cross-sectional results were replicated across two independent cohorts is a strength of the current study, and enables generalization of the findings beyond a local perspective. It also mitigates the risk that our conclusions ensue from a type I error. The cohorts differed in terms of demographics - Cohort 1 being multinational (primarily US and Israel), composed mainly of highly educated people, enriched with healthcare providers and having a high prevalence of women compared with Cohort 2, which had a larger proportion of students and young adults, was more balanced in terms of gender distribution, and composed of Israeli population exclusively. In addition, that the two studies used different measurements and still results generally replicated across studies further supports the generalizability of our findings to other populations globally and augurs well for the internal validity of the data.

In light of the urgent dilemma of lockdown versus reopening, with the tradeoff between health and economy^28^, the results of this study may have several implications for policymakers. We show here that the economic impact, which is strongly driven by lockdown policies, may have significant mental health implications like increase in depression. Due to the rising concerns regarding increased suicide risk in the face of the pandemic^29^, the possible link between the financial impact of COVID-19 and depressive symptoms may be a red flag for policy makers as they determine policies of reopening the economy. Thus, our study emphasizes the importance of maintaining balance between necessary social distancing and minimization of economic slowdown. In addition, our findings can guide clinicians as they encounter patients during the pandemic, suggesting that healthcare providers should actively probe patients for a change for the worse in their income, and ask them specifically about their subjective stress regarding the financial impact of COVID-19. Our results may suggest that these financial stressors are sensitive markers for the development of depression. Importantly, our results suggest that the impact of income loss and financial strain on depressive symptoms are independent of pre-COVID-19 income. We therefore suggest that people from all backgrounds who report stress about their financial situation during the pandemic, including those with high income, are vulnerable to the effects of the financial crisis on mental health, and therefore no one should be overlooked for their increased risk for depression.

This study had several limitations. First, the sampling was not random, rather we employed a “snowball” recruitment in both cohorts through the investigators’ social networks. Therefore, potential biases that were reported recently in online surveys during COVID-19 should be considered^27^. Second, we had substantial attrition between T1 and T2 in both studies, especially in Cohort 2, where we were likely underpowered to test longitudinal trajectories. Therefore, a type II error is possible. Third, in Cohort 1, the measure for income loss was binary, which limited us in assessing variability in the trajectory over time. Lastly, our study employed online crowdsourcing data collection, with all of its inherent limitations as participants were not interviewed in person^30^. Still, arguably even when accounting for the limitations mentioned above, that overall the association between objective (income loss) and subjective (financial worries) financial factors with depressive symptoms was replicated in two independent studies mitigates most concerns regarding generalizability of our findings.

To conclude, we provide converging evidence to suggest a specific association between financial stressors and depression during a global pandemic. A decrease in income during this time, as well as perceived financial strain, can lead to deterioration in mental health and might generate depression in a specific manner. Our findings may suggest that the “financial COVID-19” could have a serious impact on health, specifically mental health, which should be gravely considered by policy makers as they decide steps resulting in economic slowdown due to COVID-19 specific physical health concerns. In light of the growing concern about increase in suicide following the pandemic^29^, our findings provide empirical data collected during the pandemic that may help the efforts to improve early detection of and intervention with people at increased risk for depression. Future research could focus on early interventions that will enable mental health services to offer timely help to people who were financially harmed by the pandemic.

## Data Availability

Data collected for this study includes individual participants data. Data cannot be publicly accessible due to Institutional Review Board guidelines. We are open to collaborations with other researchers upon contacting us.

## Statements

## Acknowledgment

We thank participants of covid19resilience.org for their contribution to data generation. We wish to thank Amit Lewinthal, Shira Bursztyn and Dana Basel for their assistance in collecting the data. Special acknowledgment to Prof. Avner De-Shalit from the Hebrew University of Jerusalem, for his consultancy and enlightening insights.

## Statement of Ethics

The study was approved by the Institutional Review Boards of the University of Pennsylvania and Sheba Medical Center.

## Study protocols and data sharing

Data collected for this study includes individual participants’ data. Data cannot be publicly accessible due to Institutional Review Board guidelines. We are open to collaborations with other researchers upon contacting us.

## Author Contributions

All authors significantly contributed to, reviewed and approved the final manuscript.

Conceiving and designing the study: NHP, TMM, DG, ID, DMG, LAB, EV, LKW,MHH, MLS, RG, RCG, REG, IMP, RB

Data collection: NHP, GED, NM, RB Statistical analyses: TMM

Data interpretation: NHP, TMM, DG, GED, ID, DMG, LAB, NM, LKW, EV, MHH, MLS, RG, RCG, REG, IMP, RB

Writing the final manuscript: NHP, TMM, DG, IMP, RB

## References

1 Holmes EA, O’Connor RC, Perry VH, et al. Multidisciplinary research priorities for the COVID-19 pandemic: a call for action for mental health science. The Lancet Psychiatry 2020.

2 Parmet WE, Sinha MS. Covid-19—the law and limits of quarantine. N Engl J Med 2020; 382: e28.

3 Bureau of Labor Statistics. The employment situation-April 2020. 2020 https://www.bls.gov/news.release/archives/empsit_05082020.pdf.

4 World Bank. Europe and Central Asia Economic Update, Spring 2020?: Fighting COVID-19. Washington DC, 2020 http://hdl.handle.net/10986/33476.

5 World Bank. Global Economic Prospects, June 2020. Washington, DC, 2020 https://openknowledge.worldbank.org/handle/10986/33748.

6 Zimmerman FJ, Katon W. Socioeconomic status, depression disparities, and financial strain: what lies behind the income‐depression relationship? Health Econ 2005; 14: 1197–215.

7 Goldman-Mellor SJ, Saxton KB, Catalano RC. Economic contraction and mental health: A review of the evidence, 1990-2009. Int J Ment Health 2010; 39: 6–31.

8 Van Hal G. The true cost of the economic crisis on psychological well-being: a review. Psychol Res Behav Manag 2015; 8: 17.

9 Gili M, Roca M, Basu S, McKee M, Stuckler D. The mental health risks of economic crisis in Spain: evidence from primary care centres, 2006 and 2010. Eur J Public Health 2013; 23: 103–8.

10 Li J, Yang Z, Qiu H, et al. Anxiety and depression among general population in China at the peak of the COVID‐19 epidemic. World Psychiatry 2020; 19: 249.

11 Pierce M, Hope H, Ford T, et al. Mental health before and during the COVID-19 pandemic: a longitudinal probability sample survey of the UK population. The Lancet Psychiatry 2020.

12 Ettman CK, Abdalla SM, Cohen GH, Sampson L, Vivier PM, Galea S. Prevalence of Depression Symptoms in US Adults Before and During the COVID-19 Pandemic. JAMA Netw Open 2020; 3: e2019686–e2019686.

13 Shanahan L, Steinhoff A, Bechtiger L, et al. Emotional distress in young adults during the COVID-19 pandemic: evidence of risk and resilience from a longitudinal cohort study. Psychol Med 2020; : 1–32.

14 Sanchez DG, Parra NG, Ozden C, Rijkers B. Which Jobs Are Most Vulnerable to COVID-19? What an Analysis of the European Union Reveals. Res Policy Br 2020.

15 McKee M, Stuckler D. If the world fails to protect the economy, COVID-19 will damage health not just now but also in the future. Nat Med 2020; 26: 640–2.

16 Czeisler MÉ. Mental Health, Substance Use, and Suicidal Ideation During the COVID-19 Pandemic—United States, June 24–30, 2020. MMWR Morb Mortal Wkly Rep 2020; 69.

17 Barzilay R, Moore TM, Greenberg DM, et al. Resilience, COVID-19-related stress, anxiety and depression during the pandemic in a large population enriched for healthcare providers. Transl Psychiatry 2020.

18 Wilkinson LR. Financial strain and mental health among older adults during the great recession. Journals Gerontol Ser B Psychol Sci Soc Sci 2016; 71: 745–54.

19 Spitzer RL, Kroenke K, Williams JBW, Löwe B. A brief measure for assessing generalized anxiety disorder: the GAD-7. Arch Intern Med 2006; 166: 1092–7.

20 Arroll B, Goodyear-Smith F, Crengle S, et al. Validation of PHQ-2 and PHQ-9 to screen for major depression in the primary care population. Ann Fam Med 2010; 8: 348–53.

21 Kuznetsova A, Brockhoff PB, Christensen RHB. lmerTest Package: Tests in Linear Mixed Effects Models. J Stat Softw 2017; 82. DOI:10.18637/jss.v082.i13.

22 Bevans M, Ross A, Cella D. Patient-Reported Outcomes Measurement Information System (PROMIS): efficient, standardized tools to measure self-reported health and quality of life. Nurs Outlook 2014; 62: 339–45.

23 Mosheva M, Hertz-Palmor N, Dorman Ilan S, et al. Anxiety, pandemic-related stress and resilience among physicians during the COVID-19 pandemic. Depress Anxiety 2020; published online Aug. DOI:10.1002/da.23085.

24 Imai H, Matsuishi K, Ito A, et al. Factors associated with motivation and hesitation to work among health professionals during a public crisis: a cross sectional study of hospital workers in Japan during the pandemic (H1N1) 2009. BMC Public Health 2010; 10: 672.

25 Economou M, Madianos M, Peppou LE, Patelakis A, Stefanis CN. Major depression in the era of economic crisis: a replication of a cross-sectional study across Greece. J Affect Disord 2013; 145: 308–14.

26 Selenko E, Batinic B. Beyond debt. A moderator analysis of the relationship between perceived financial strain and mental health. Soc Sci Med 2011; 73: 1725–32.

27 Pierce M, McManus S, Jessop C, et al. Says who? The significance of sampling in mental health surveys during COVID-19. The Lancet Psychiatry 2020.

28 Covid-19 hurts the most vulnerable – but so does lockdown. We need more nuanced debate. Guard. 2020. https://www.theguardian.com/commentisfree/2020/may/16/covid-19-coronavirus-lockdown-economy-debate.

29 Reger MA, Stanley IH, Joiner TE. Suicide mortality and coronavirus disease 2019—a perfect storm? JAMA psychiatry 2020.

30 Behrend TS, Sharek DJ, Meade AW, Wiebe EN. The viability of crowdsourcing for survey research. Behav Res Methods 2011; 43: 800.

